# Pulmonary Function Outcomes after Tuberculosis treatment in Children: A Systematic Review and Meta-analysis

**DOI:** 10.1101/2023.10.13.23297035

**Authors:** Yao Long Lew, Angelica F Tan, Stephanie T Yerkovich, Tsin Wen Yeo, Anne B Chang, Chris Lowbridge

**Author notes:** **Corresponding author** details Yao Long Lew, Child and Maternal Health Division, Menzies School of Health Research, John Matthews Building, Royal Darwin Hospital Campus, Rocklands Drive, Tiwi, NT 0810, Australia.

## Abstract

**Background:** Despite tuberculosis (TB) being a curable disease, current guidelines fail to account for long-term outcomes of post-tuberculosis lung disease (PTLD) – a cause of global morbidity despite successful completion of effective treatment. Our systematic review aimed to synthesise the available evidence on the lung function outcomes of childhood pulmonary tuberculosis (PTB).

**Methods:** PubMed, ISI Web of Science, the Cochrane Library, and ProQuest databases were searched for English-only studies without time restriction (latest search date 22 March 2023). Inclusion criteria were (1) patients who had TB with pulmonary involvement at age ≤18 years; (2) pulmonary function tests (PFTs) performed on patients after treatment completion; and (3) observational studies, including cohort and cross-sectional studies. We adhered to the recommendations of the Cochrane Collaboration and the Preferred Reporting Items for Systematic Reviews and Meta-Analyses (PRISMA).

**Results:** From 8040 records, five studies were included (involving n=567 children) with spirometry measures from four studies included into meta-analyses. The effect size of childhood TB on forced expiratory volume in the first second (FEV_1_) and forced vital capacity (FVC) z-scores were estimated to be -1.53 (95% CI: -2.65, -0.41; p=0.007) and -1.93 (95% CI: -3.35, -0.50; p=0.008) respectively.

**Discussion:** The small number of included studies reflects this under-researched area, relative to the global burden of TB. Nevertheless, as childhood PTB impacts future lung function, pulmonary function tests (such as spirometry) should be considered a routine test when evaluating the long-term lung health of children beyond their completion of TB treatment.

**What is already known on this topic?:** Tuberculosis (TB) is a treatable disease, but despite resolution of the infection, lung function deficits associated with post-tuberculosis lung disease (PTLD) can persist. While this is well-appreciated in adults, the extent and severity of PTLD in children is not well characterised. This area of work is important because of the potential long-term impacts of PTLD on children’s lung health and development.

**What this study adds:** Our meta-analyses showed that childhood TB causes significant decline in at least two spirometry parameters despite high levels of between-study heterogeneity. The effect size of childhood TB on forced expiratory volume in the first second (FEV_1_) and forced vital capacity (FVC) z-scores were estimated to be -1.53 (95% CI: -2.65, -0.41; p=0.007) and -1.93 (95% CI: -3.35, -0.50; p=0.008). A previous meta-analysis of spirometric data from adult populations with drug-susceptible TB gave combined estimated mean of 76.6% (95% CI: 71.6, 81.6) and 81.8% (95% CI: 77.4, 86.2) of predicted FEV_1_ and FVC respectively. While direct comparison with this current study was not possible, it suggests that childhood TB results in lung function decline just as much as adult TB, if not more so.

**How this study might affect research, practice, or policy:** This study supports incorporation of routine pulmonary function tests into the follow-ups of children with prior history of TB, allowing for early detection and management of PTLD.

## INTRODUCTION

Tuberculosis (TB) is an airborne disease caused by Mycobacterium tuberculosis. Inhalation of the bacterium into airways can result in TB infection. Pulmonary disease is typically established when the host’s innate immune response is unable to eliminate the bacterium.^1^ In 2021, an estimated 10.6 million people fell ill from TB globally, children under 15 years old accounted for 11% of this burden.^2^ Childhood TB causes a spectrum of clinical presentations, most commonly pulmonary disease. Irrespective of organ involvement, obtaining bacteriological confirmation for infants and young children still proves challenging. Age is the key determinant of disease progression, with risk of progression to pulmonary tuberculosis (PTB) about 30-40% when primary infection occurs in infants under a year old.^3^ While improving diagnosis and management of childhood TB is important,^4^ children with prior PTB can experience detrimental changes irrespective of successful completion of treatment.^5^ There is a significant knowledge gap in the occurrence and severity of post-tuberculosis lung disease (PTLD) in children.^6^ PTLD in adults is better described, this includes post-TB bronchiectasis^7^ and changes in lung function.^8^ One study reported decline in mean FEV_1_ and FVC z-scores by -1.07 and -0.91 upon treatment completion, and -0.91 and -0.64 respectively three years post-treatment.^9^

Specific data on childhood PTLD is required as early-life lung injuries from respiratory infections and pneumonia cause deficits during children’s peak lung growth and development.^10^ It is possible that childhood PTB will be more detrimental to future lung function, compared to acquiring PTB as TB-naïve adults. This was highlighted in a recent review whereby the authors recommended evaluation of PTLD beyond completion of routine treatment using objective tests for early detection of post-TB pulmonary changes irrespective of symptoms.^11^ These could promote initiation of treatments that may prevent irreversible lung function decline, reduce healthcare costs, and alleviate burden to patients, their families, and healthcare systems.

Given the absence of a systematic review evaluating effects of childhood TB on pulmonary function tests (PFTs) outcomes, we undertook this review and meta-analysis aiming to synthesise available evidence regarding effects of childhood PTB on future lung function.

## METHODS

Review and analysis findings were reported in accordance to the Preferred Reporting Items for Systematic Review and Meta-Analyses (PRISMA) guidelines and checklist.^12^

### Literature Search

A search strategy was developed to search the PubMed, Cochrane Library, and ISI Web of Science databases for eligible studies up to 22 March 2023 (appendix I). Grey literature searches were performed on ProQuest database, followed by manual citation searching of included studies. No publications were excluded based on publication date.

### Eligibility criteria

Studies which fulfilled the following inclusion criteria were: (1) patients who had TB with pulmonary involvement at age ≤18 years; (2) PFTs performed on patients after treatment completion; and (3) observational studies, including cohort and cross-sectional studies. Exclusion criteria were as follows: (1) mixed-population studies which did not report the ≤18 years subgroup separately; (2) TB studies without pulmonary involvement; (3) evidence of non-standard anti-TB treatment regimens; (4) did not perform PFTs after treatment; or (5) reviews and case studies.

Studies from all countries and settings were included. Studies were included regardless of bacteriological confirmation, unreported treatment regimens, or timing of PFTs. Studies with other concurrent disease as primary domain were included providing that PFT measures were sufficiently reported for inclusion in our analysis.

### Outcome

The primary outcomes were spirometry results. The secondary outcomes were measurements from non-spirometry PFTs. There were no limits on timing of PFTs after completion of TB treatment.

### Data extraction and quality assessment

Screening and eligibility assessment was performed by two reviewers independently (YLL and AFT). References from eligible studies were assessed to ensure inclusion of relevant studies. No automation tools were used throughout the review process. For eligible studies, data extraction was performed according to a standardised collection form (appendix II). Using the Newcastle-Ottawa Scale, both reviewers independently assessed the studies’ risk of bias and certainty assessments, consensus was achieved through discussion.^13^

### Statistical analysis

All statistical analysis was done using R for Windows, version 4.2.2.^14^ Median and interquartile range were normalised to give mean and standard deviation where distance between Q1-to-median was equal to median-to-Q3.^15^ Spirometry results presented as percentage of predicted values were converted to z-scores using the “rspiro” package,^16^ based on GLI-2012 equation^17^ accounting for North East Asian ethnicity, mean age 11.9 years, a male-to-female ratio of 53.5:46.5, and median height-for-age based on 2017 Korean National Growth Charts.^18^ Studies which reported primary outcome measures were included in meta-analyses, effect sizes were calculated using Hedges’ g and presented with 95% confidence intervals (CI).^19^ Outcome measures not included in meta-analyses were presented in separate tables with relevant summary statistics.

We used random effects models (DerSimonian-Laird method) to estimate overall effect due to variable data with significant heterogeneity. Between-study heterogeneity was assessed using the I^2^-statistic, with values >75% representing considerable heterogeneity. Meta-analyses were performed using “metafor” package.^20^ Sensitivity analysis was not performed as substantial inter-study differences rendered statistical approaches meaningless.

## RESULTS

### Search results

The screening process is detailed in Figure 1. After removing duplicates, 8040 records were screened, from which we reviewed 34 full-text articles, and finally included five studies.^21-25^ Characteristics of excluded studies were summarised in appendix III.

**Figure 1:**
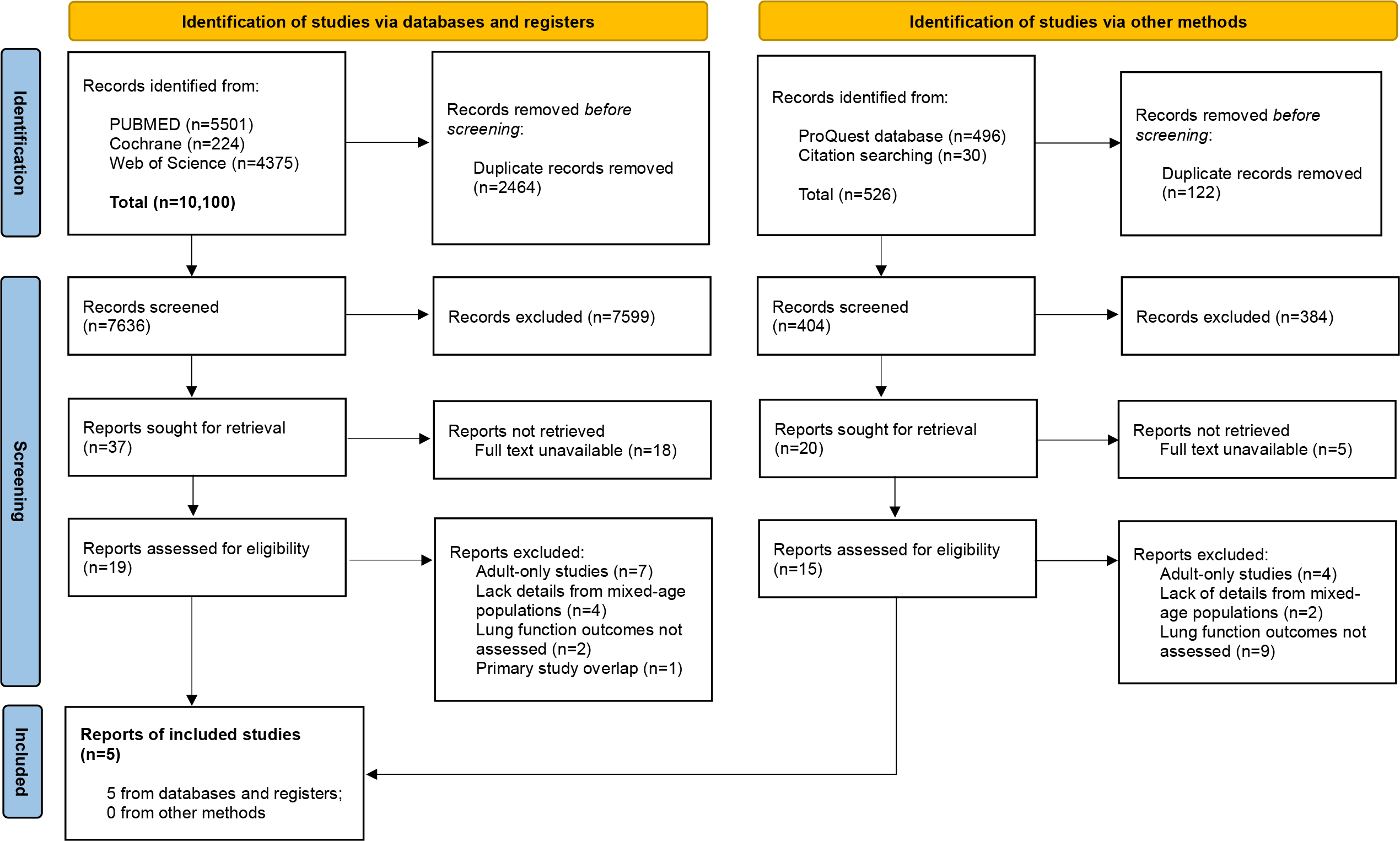
PRISMA diagram showing identification, screening, and inclusion of studies.

Included studies were cohort or cross-sectional studies conducted in urban or peri-urban settings of TB-endemic countries in Africa, except for one retrospective review study in South Korea that has an upper-moderate TB incidence.^22^ A total of 567 children with history of PTB were included; median number of children in studies was 68 (range: 42-305).^21-25^ Key characteristics were reported in Table 1, and details of quality scores shown in appendix IV. One study performed non-spirometry PFTs after early-life TB,^25^ thus excluded from meta-analyses. Granular details of studies included in meta-analyses are provided in appendix V.

**Table 1:**
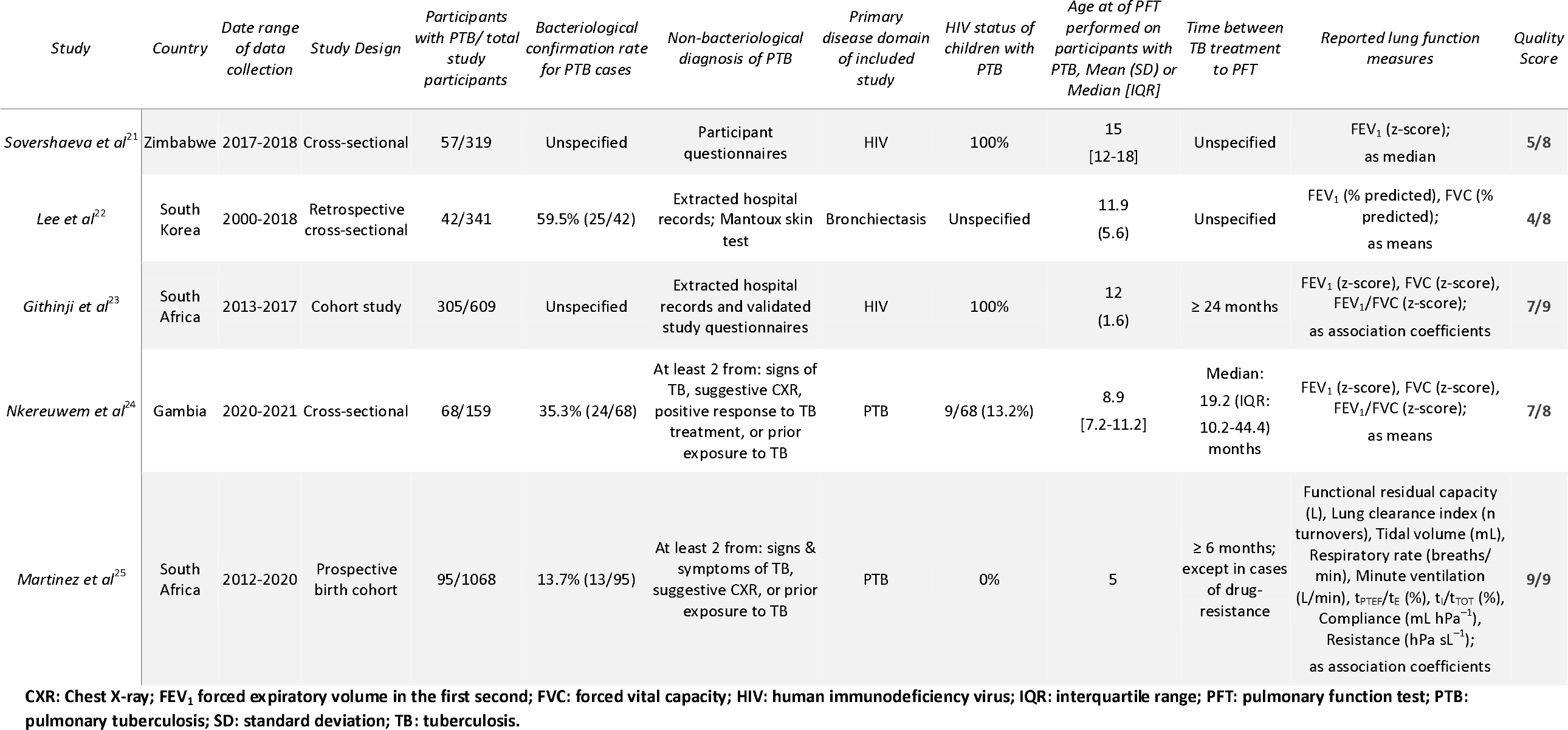
Included studies and their characteristics.

| Study | Country | Date range of data collection | Study Design | Participants with PTB/ total study participants | Bacteriological confirmation rate for PTB cases | Non-bacteriological diagnosis of PTB | Primary disease domain of included study | HIV status of children with PTB | Age at of PFT performed on participants with PTB, Mean (SD) or Median [IQR] | Time between TB treatment to PFT | Reported lung function measures | Quality Score |
| --- | --- | --- | --- | --- | --- | --- | --- | --- | --- | --- | --- | --- |
| Sovershaeva et al<sup>21</sup> | Zimbabwe | 2017-2018 | Cross-sectional | 57/319 | Unspecified | Participant questionnaires | HIV | 100% | 15 [12-18] | Unspecified | FEV<sub>1</sub> (z-score); as median | 5/8 |
| Lee et al<sup>22</sup> | South Korea | 2000-2018 | Retrospective cross-sectional | 42/341 | 59.5% (25/42) | Extracted hospital records; Mantoux skin test | Bronchiectasis | Unspecified | 11.9 (5.6) | Unspecified | FEV<sub>1</sub> (% predicted), FVC (% predicted); as means | 4/8 |
| Githinji et al<sup>23</sup> | South Africa | 2013-2017 | Cohort study | 305/609 | Unspecified | Extracted hospital records and validated study questionnaires | HIV | 100% | 12 (1.6) | ≥ 24 months | FEV<sub>1</sub> (z-score), FVC (z-score), FEV<sub>1</sub>/FVC (z-score); as association coefficients | 7/9 |
| Nkereuwem et al<sup>24</sup> | Gambia | 2020-2021 | Cross-sectional | 68/159 | 35.3% (24/68) | At least 2 from: signs of TB, suggestive CXR, positive response to TB treatment, or prior exposure to TB | PTB | 9/68 (13.2%) | 8.9 [7.2-11.2] | Median: 19.2 (IQR: 10.2-44.4) months | FEV<sub>1</sub> (z-score), FVC (z-score), FEV<sub>1</sub>/FVC (z-score); as means | 7/8 |
| Martinez et al<sup>25</sup> | South Africa | 2012-2020 | Prospective birth cohort | 95/1068 | 13.7% (13/95) | At least 2 from: signs & symptoms of TB, suggestive CXR, or prior exposure to TB | PTB | 0% | 5 | ≥ 6 months; except in cases of drug-resistance | Functional residual capacity (L), Lung clearance index (n turnovers), Tidal volume (mL), Respiratory rate (breaths/min), Minute ventilation (L/min), t<sub>PTEF</sub>/t<sub>E</sub> (%), t<sub>I</sub>/t<sub>TOT</sub> (%), Compliance (mL hPa<sup>-1</sup>), Resistance (hPa sL<sup>-1</sup>); as association coefficients | 9/9 |

CXR: Chest X-ray; FEV<sub>1</sub> forced expiratory volume in the first second; FVC: forced vital capacity; HIV: human immunodeficiency virus; IQR: interquartile range; PFT: pulmonary function test; PTB: pulmonary tuberculosis; SD: standard deviation; TB: tuberculosis.

#### Diagnosis and treatment of PTB

Bacteriological confirmation of TB varied between studies, ranging from 13.7%^25^ to 58.5%.^22^ GeneXpert MTB/RIF® was used to rule out active infection pre-spirometry.^21 26^ Only one study reported completion of treatment regimen for drug-susceptible TB according to national guidelines, modified for drug-resistance;^25^ other studies^21-24^ did not report treatment regimen details.

#### Pulmonary function tests

The time between treatment completion to PFTs ranged between 6 to 24 months in the three most recent studies;^23-25^ two earlier studies^21 22^ did not specify this duration. Three studies^21-23^ reported performing spirometry according to American Thoracic Society/European Respiratory Society 2005 standards,^27^ and one^24^ according to the 2019 update.^28^ Three studies^21 23 24^ performed bronchodilator responsiveness testing.

For effects of childhood TB on FEV_1_ and FVC z-scores, meta-analyses were possible and presented in Forest plots with pooled effect size estimates of -1.53 (95% CI: -2.65, -0.41; p=0.007; Figure 2) and - 1.93 (95% CI: -3.35, -0.50; p=0.008; Figure 3).

**Figure 2:**
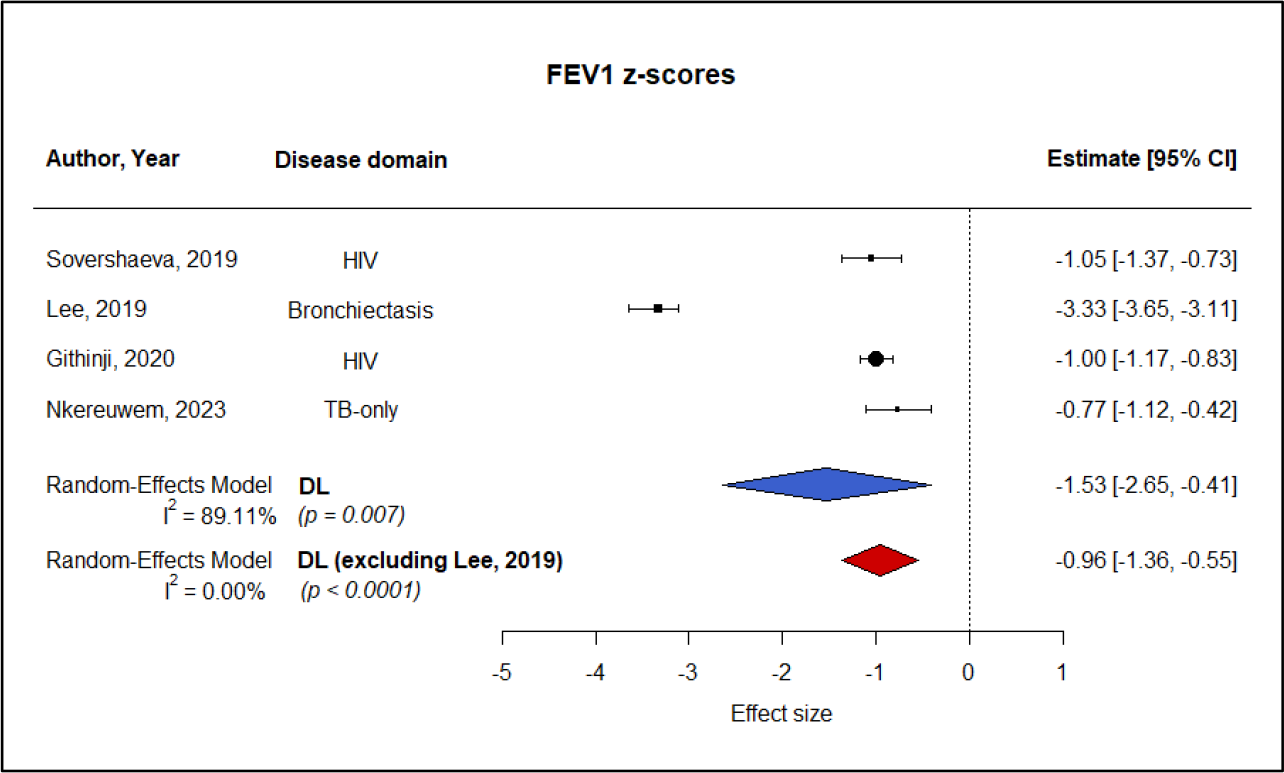
Forest plot of effect sizes of childhood PTB on FEV_1_ z-scores.

**Figure 3:**
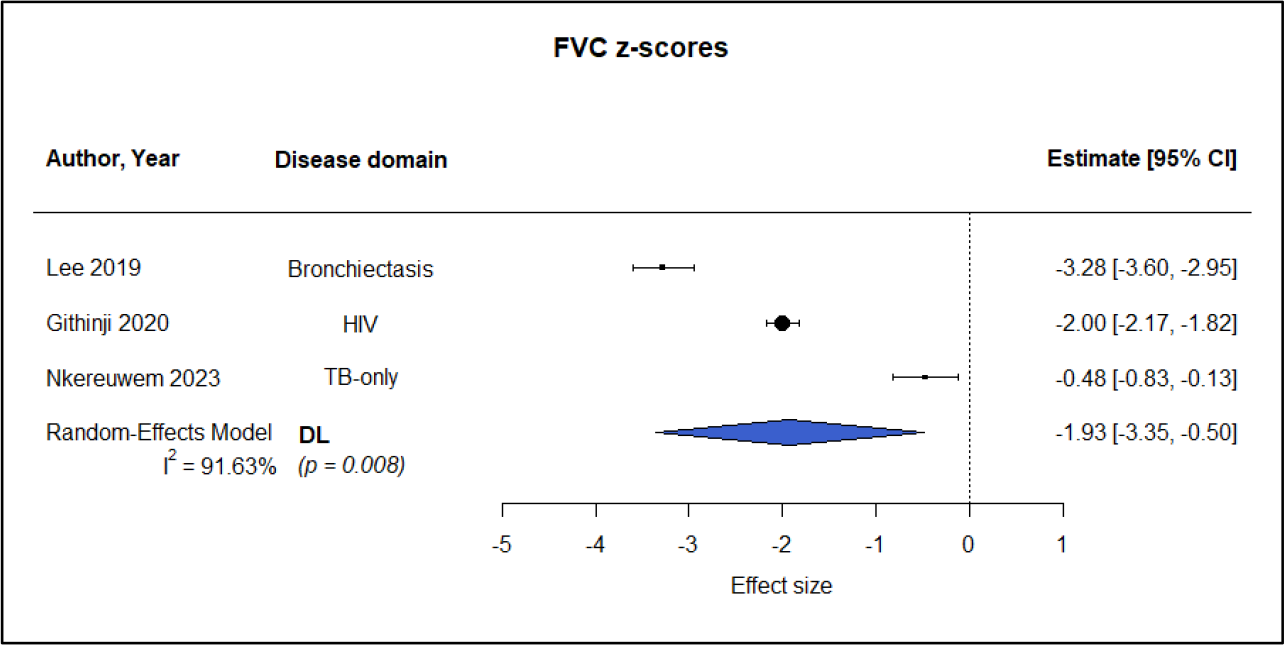
Forest plot of effect sizes of childhood PTB on FVC z-scores.

Meta-analysis was not possible for FEV_1_/FVC ratios presented in included studies, thus summarised in Table 2 instead. Only one included study performed non-spirometry PFTs, they instead reported association coefficients between childhood PTB occurring between one to four years of age with measurements taken at the age of five, as presented in appendix VI.^25^

**Table 2:** Summary of studies reporting FEV_1_/FVC ratios in any manner.

| Study | Disease domain | Metrics | Mean | Association coefficient | P-values | Details |
| --- | --- | --- | --- | --- | --- | --- |
| Lee *et al*<sup>22</sup> | Bronchiectasis + TB | % predicted | Plotted <sup>a</sup> | n.a. | n.a. | <sup>a</sup> mean FEV<sub>1</sub>/FVC ratio was presented as an interval plot but value provided was incorrectly labelled |
| Githinji *et al*<sup>23</sup> | HIV + PTB | z-scores | n.a. | -0.01 <sup>b</sup> | 0.904 | <sup>b</sup> 95% CI unreported |
| Nkereuwem *et al*<sup>24</sup> | PTB-only | z-scores | -0.54 (0.91) | n.a. | 0.001 |  |

#### Heterogeneity, Sensitivity, and Bias

Due to the small number of studies, sensitivity analysis was not performed. Significant between-study heterogeneity was observed in meta-analyses for FEV_1_ and FVC effect sizes at I^2^=89.11% (p<0.0001) and I^2^=91.63% (p<0.0001) respectively, as indicated by blue diamonds. The study^22^ which reported PTB-bronchiectasis overlap had the largest effect size on both FEV_1_ and FVC z-scores. The exclusion of this study^22^ from the meta-analysis for FEV_1_ led to significant reduction in heterogeneity as shown by red diamond in Figure 2, but not observed in the meta-analysis for FVC.

Publication bias was judged as unlikely as included studies were observational, funded by research grants, and unlikely to be influenced by industry-based sponsorship or agenda. It was notable that three included studies reported primarily on non-TB diseases, thus less likely to be affected by reporting bias in terms of PTB-related outcomes, at the cost of being less comprehensive in reporting PTB-related details.^21-23^ One included study had significant information bias as summary statistics were not reported numerically, necessitating pixel-counting of error bars from interval plots to approximate the data dispersion within PTB subgroup.^22^

## DISCUSSION

### Interpretation of results

The small number of included studies highlights under-representation of childhood TB globally.^2^ The overall direction of the effects of PTB on lung function were negative, i.e., reduced lung function in both meta-analyses of FEV_1_ and FVC. These findings align with the current understanding of PTLD in adults,^29^ which lends support to the validity of our approach. While pooled effect sizes appear to be significant, high I^2^ values indicate substantial between-study heterogeneity which is a key limitation in our study. This suggests a research gap in quantifying the impact of PTB during childhood on lung function outcomes, particularly in high-prevalence settings.

Of the included studies, three had primary diseases^21-23^ that were not PTB, which were reasonable to include as HIV-coinfection is a significant comorbidity,^30^ and bronchiectasis is a well-established sequela of PTB.^6^ One study^24^ evaluated health-related quality of life post-PTB, suggestive of recent paradigm shifts to better evaluate PTLD. As only two studies reported spirometry performed at >24 months^23^ and >6 months^24^ after treatment completion, the actual effect of spirometry timings was indeterminate due to variability of included studies. A prospective cohort of adult TB survivors did show greater deterioration in FEV_1_ and FVC values three years after treatment completion^9^ compared to the first year post-treatment^29^.

Due to a low quality score, contextual interpretation for one included study^22^ is presented here. As the GLI-2012 equation^17^ for North East Asian ethnicity was developed using only subjects aged ≥16 years, spirometry z-scores for this study^22^ were calculated using extrapolation for young children, which may have inadvertently inflated effects sizes. This inference was partially supported by a validation study^31^ which found that South Korean females aged 7-8 years have mean FEV_1_ and FVC z-scores lower than GLI-2012 predictions by -0.23 (95% CI: -0.31, -0.15) and -0.26 (95% CI: -0.36, -0.16) respectively, suggesting actual lung function for this subgroup is slightly below established baseline. A secondary analysis which excluded this outlier study^22^ from meta-analyses yielded an alternative random-effects model for FEV_1_ based on three studies with reduced statistical heterogeneity (red diamond; Figure 2). As pooled effect sizes regardless of exclusion remained below -0.8, overall interpretation was that childhood TB exerts a large effect^32^ on FEV_1_ . Removal of this study^22^ did not appreciably change the pooled effect size estimate nor the I^2^-statistic for FVC (not shown). It is noteworthy that one study^23^ reported more significant HIV-associated decline in FVC than FEV_1_ ; the combinatory effect of PTB within an all-HIV cohort gave a greater change in FVC relative to baseline, and subsequently a larger standardised effect size as compared to FEV_1_. This was partly supported by another study^10^ which found early childhood respiratory infections had a marginally greater effect on FVC than FEV_1_, raising plausibility that HIV-coinfection could be a clinical contributor to heterogeneity observed in Figure 3.

### Limitations of evidence and review process

The included evidence had limitations inherent to population, nature of disease, and outcome measures. The World Health Organization (WHO) classifies childhood TB as diagnosed in children <15 years old, leading to bias in age stratification at study design level.^2^ Adolescents aged ≥15 years are classified as adults, inadvertently excluding evidence encompassing full age range of childhood. Bacteriological confirmation of TB was relatively low (range: 13.7-59.5%), thus misclassification bias among children whose TB was diagnosed clinically is possible.^33^ One study^22^ did not report numerical values for standard deviations thus pixel-counting from published figures was performed, measurement errors may be propagated when calculating Hedge’s g. One study^23^ reported z-score changes as association coefficients thus necessitating Fisher’s z-transformation,^34^ resulting in confidence intervals that were much smaller than all other studies^21 22 24^ included in meta-analyses. Thus, the pooled effect sizes should be interpreted with awareness of our approach used.

### Clinical and Policy Implications

To the best of our knowledge, this is the first meta-analysis to investigate effects of childhood PTB on lung function decline. Our findings suggest that childhood PTB is associated with overall decreases in subsequent FEV_1_ and FVC z-scores. Concurrent bronchiectasis exerted the greatest additive negative impact on spirometry parameters compared to HIV-coinfection or TB on its own.^22^ Childhood TB and resultant PTLD remain understudied within paediatric populations despite clear association with lung function decline in children and adolescents, further compounded by underdiagnosis and subsequent failure to treat. ^2 6^

WHO-defined outcomes of TB treatment include cured or treatment completed positive outcomes, and negative outcomes of lost to follow-up, treatment failure, or death.^35^ These outcome indicators are based primarily on bacteriological clearance and treatment compliance, with post-TB sequelae and residual respiratory impairment unaccounted for. The most recent roadmap for ending TB in children and adolescents does not address the fact that post-TB disabilities and PTLD do occur beyond completion of treatment.^36^

In the first consensus-based set of clinical standards for PTLD,^37^ the foremost standard recommends clinical, functional, and subjective evaluation of every patient completing TB treatment for PTLD, with considerations for paediatric care including selection of age-appropriate PFTs and quality-of-life questionnaires. The second and third standards called for evaluating patients with PTLD for pulmonary rehabilitation (PR), and the organisation of PR programmes with health settings and individual patient’s needs in mind. While not routinely done for children and thus far unreported for childhood PTB, individualised PR programmes have been attempted for paediatric asthma^38^ and could be adjusted younger patients in high-TB settings.

Objective lung function measurements allow for prompt initiation of PR^37^ or other adjunctive therapies^39^ to prevent late-life onset of respiratory diseases such as bronchiectasis, asthma, or chronic obstructive pulmonary disease. As performing spirometry on young children can be challenging, non-spirometry PFTs should be considered for children below certain ages and others on a case-by-case basis. At least one study has explored oscillometry for children above two years, alongside spirometry for those above four years of age.^40^ Subsequent findings may address the evidence gap for performing scheduled PFTs as part of national TB programmes or routine post-TB pulmonary health surveillance^11^, especially in low-to-middle income countries with significant disease burden.

Our findings suggest that spirometry or other PFTs should be performed as routine follow-up of children beyond TB treatment completion to evaluate their lung function and diagnose impaired lung function. Lung health monitoring would enable appropriate and timely interventions to reduce the frequency and severity of PTLD beyond treatment completion.

### Other information

This systematic review was registered in PROSPERO under the registration number: CRD42021250172. Two deviations from registered protocol were as follows: Firstly, redefining primary outcomes as lung function measured using spirometry and secondary outcomes as lung function by non-spirometry PFTs, as other published reviews have evaluated non-PFT secondary outcomes. Secondly, number of search databases were reduced due to duplication of records.

YLL is supported via the Malaysia Australia Colombo Plan Commemoration (MACC) and Australian Government Research Training Program (RTP) Scholarship at Charles Darwin University. All other authors do not declare any financial support or sponsorship for this study.

There are no competing interests to declare.

## Data Availability

All data relevant to the study are included in the article or uploaded as supplementary information

## REFERENCES

1. Cohen SB, Gern BH, Delahaye JL, et al. Alveolar Macrophages Provide an Early Mycobacterium tuberculosis Niche and Initiate Dissemination. Cell Host Microbe 2018;24(3):439–46 e4. doi: 10.1016/j.chom.2018.08.001 [published Online First: 2018/08/28]

2. World Health Organization. Global tuberculosis report 2022. Geneva 2022.

3. Marais BJ, Gie RP, Schaaf HS, et al. The natural history of childhood intra-thoracic tuberculosis: a critical review of literature from the pre-chemotherapy era. Int J Tuberc Lung Dis 2004;8(4):392–402. [published Online First: 2004/05/15]

4. Howard-Jones AR, Marais BJ. Tuberculosis in children: screening, diagnosis and management. Curr Opin Pediatr 2020;32(3):395–404. doi: 10.1097/mop.0000000000000897 [published Online First: 2020/05/07]

5. Gohar Ali M, Syed Muhammad Z, Shahzad T, et al. Post tuberculosis sequelae in patients treated for tuberculosis: An observational study at a tertiary care center of a high TB burden country. European Respiratory Journal 2018;52(suppl 62):PA2745. doi: 10.1183/13993003.congress-2018.PA2745

6. Allwood BW, Byrne A, Meghji J, et al. Post-Tuberculosis Lung Disease: Clinical Review of an Under-Recognised Global Challenge. Respiration 2021;100(8):751–63. doi: 10.1159/000512531 [published Online First: 2021/01/06]

7. Chalmers JD, Chang AB, Chotirmall SH, et al. Bronchiectasis. Nat Rev Dis Primers 2018;4(1):45. doi: 10.1038/s41572-018-0042-3 [published Online First: 2018/11/18]

8. Ivanova O, Hoffmann VS, Lange C, et al. Post-tuberculosis lung impairment: systematic review and meta-analysis of spirometry data from 14 621 people. European Respiratory Review 2023;32(168)

9. Nightingale R, Chinoko B, Lesosky M, et al. Respiratory symptoms and lung function in patients treated for pulmonary tuberculosis in Malawi: a prospective cohort study. Thorax 2022;77(11):1131–39. doi: 10.1136/thoraxjnl-2021-217190

10. Collaro AJ, McElrea MS, Marchant JM, et al. The effect of early childhood respiratory infections and pneumonia on lifelong lung function: a systematic review. The Lancet Child & Adolescent Health 2023 doi: 10.1016/S2352-4642(23)00030-5 [published Online First: 2023/04/11]

11. Nkereuwem E, Togun T, Kampmann B. Making a case for investing in post-tuberculosis lung health in children. Lancet Respir Med 2022;10(6):536–37. doi: 10.1016/S2213-2600(22)00102-3 [published Online First: 2022/03/27]

12. Page MJ, Moher D, Bossuyt PM, et al. PRISMA 2020 explanation and elaboration: updated guidance and exemplars for reporting systematic reviews. bmj 2021;372

13. Peterson J, Welch V, Losos M, et al. The Newcastle-Ottawa scale (NOS) for assessing the quality of nonrandomised studies in meta-analyses. Ottawa: Ottawa Hospital Research Institute 2011;2(1):1–12.

14. R Core Team. R: A language and environment for statistical computing. In: Computing RFfS, ed. Vienna, Austria: https://www.R-project.org/, 2022.

15. Higgins J, Green S. Medians and interquartile ranges. Cochrane Handbook for Systematic Reviews of Interventions Version;5(0)

16. Lytras T. rspiro: Implementation of Spirometry Equations: https://CRAN.R-project.org/package=rspiro, 2020.

17. Quanjer PH, Stanojevic S, Cole TJ, et al. Multi-ethnic reference values for spirometry for the 3-95-yr age range: the global lung function 2012 equations. Eur Respir J 2012;40(6):1324–43. doi: 10.1183/09031936.00080312 [published Online First: 2012/06/30]

18. Kim JH, Yun S, Hwang SS, et al. The 2017 Korean National Growth Charts for children and adolescents: development, improvement, and prospects. Korean J Pediatr 2018;61(5):135–49. doi: 10.3345/kjp.2018.61.5.135 [published Online First: 2018/06/02]

19. Hedges LV, Olkin I. Statistical methods for meta-analysis: Academic press 2014.

20. Viechtbauer W. Conducting meta-analyses in R with the metafor package. Journal of statistical software 2010;36(3):1–48.

21. Sovershaeva E, Kranzer K, McHugh G, et al. History of tuberculosis is associated with lower exhaled nitric oxide levels in HIV-infected children. Aids 2019;33(11):1711–18. doi: 10.1097/qad.0000000000002265 [published Online First: 2019/05/21]

22. Lee E, Shim JY, Kim HY, et al. Clinical characteristics and etiologies of bronchiectasis in Korean children: A multicenter retrospective study. Respir Med 2019;150:8–14. doi: 10.1016/j.rmed.2019.01.018 [published Online First: 2019/04/10]

23. Githinji LN GD, Hlengwa S, Machemedze T, Zar HJ. Longitudinal Changes in Spirometry in South African Adolescents Perinatally Infected With Human Immunodeficiency Virus Who Are Receiving Antiretroviral Therapy. Clinical infectious diseases : an official publication of the Infectious Diseases Society of America 2020;70(3):483–90. doi: doi:

24. Nkereuwem E, Agbla S, Sallahdeen A, et al. Reduced lung function and health-related quality of life after treatment for pulmonary tuberculosis in Gambian children: a cross-sectional comparative study. Thorax 2023;78(3):281–87. doi: 10.1136/thorax-2022-219085

25. Martinez L, Gray DM, Botha M, et al. The Long-Term Impact of Early-Life Tuberculosis Disease on Child Health: A Prospective Birth Cohort Study. Am J Respir Crit Care Med 2023;0(ja):null. doi: 10.1164/rccm.202208-1543OC [published Online First: 2023/02/07]

26. Gonzalez-Martinez C, Kranzer K, McHugh G, et al. Azithromycin versus placebo for the treatment of HIV-associated chronic lung disease in children and adolescents (BREATHE trial): study protocol for a randomised controlled trial. Trials 2017;18(1):622. doi: 10.1186/s13063-017-2344-2

27. Miller MR, Hankinson J, Brusasco V, et al. Standardisation of spirometry. Eur Respir J 2005;26(2):319–38. doi: 10.1183/09031936.05.00034805 [published Online First: 2005/08/02]

28. Graham BL, Steenbruggen I, Miller MR, et al. Standardization of Spirometry 2019 Update. An Official American Thoracic Society and European Respiratory Society Technical Statement. Am J Respir Crit Care Med 2019;200(8):e70–e88. doi: 10.1164/rccm.201908-1590ST [published Online First: 2019/10/16]

29. Meghji J, Lesosky M, Joekes E, et al. Patient outcomes associated with post-tuberculosis lung damage in Malawi: a prospective cohort study. Thorax 2020;75(3):269–78. doi: 10.1136/thoraxjnl-2019-213808 [published Online First: 2020/02/28]

30. Tiberi S, Carvalho ACC, Sulis G, et al. The cursed duet today: Tuberculosis and HIV-coinfection. La Presse Médicale 2017;46(2):e23–e39.

31. Park JS, Suh DI, Choi YJ, et al. Pulmonary function of healthy Korean children from three independent birth cohorts: Validation of the Global Lung Function Initiative 2012 equation. Pediatr Pulmonol 2021;56(10):3310–20. doi: 10.1002/ppul.25622 [published Online First: 2021/08/11]

32. Cohen J. Statistical power analysis for the behavioral sciences: Academic press 2013.

33. Graham SM, Cuevas LE, Jean-Philippe P, et al. Clinical Case Definitions for Classification of Intrathoracic Tuberculosis in Children: An Update. Clinical Infectious Diseases 2015;61(suppl_3):S179–S87. doi: 10.1093/cid/civ581

34. Fisher RA. Frequency distribution of the values of the correlation coefficient in samples from an indefinitely large population. Biometrika 1915;10(4):507–21.

35. Linh NN, Viney K, Gegia M, et al. World Health Organization treatment outcome definitions for tuberculosis: 2021 update. Eur Respir J 2021;58(2):2100804. doi: 10.1183/13993003.00804-2021 [published Online First: 2021/08/21]

36. World Health Organization. Roadmap towards ending TB in children and adolescents. 2018 37.

37. Migliori GB, Marx FM, Ambrosino N, et al. Clinical standards for the assessment, management and rehabilitation of post-TB lung disease. International Journal of Tuberculosis and Lung Disease 2021;25(10):797–813.

38. Kirkby S, Rossetti A, Hayes D, Jr., et al. Benefits of pulmonary rehabilitation in pediatric asthma. Pediatr Pulmonol 2018;53(8):1014–17. doi: 10.1002/ppul.24041 [published Online First: 2018/05/08]

39. Wallis RS, Ginindza S, Beattie T, et al. Adjunctive host-directed therapies for pulmonary tuberculosis: a prospective, open-label, phase 2, randomised controlled trial. The Lancet Respiratory medicine 2021;9(8):897–908.

40. Dewandel I, van Niekerk M, Ghimenton-Walters E, et al. UMOYA: a prospective longitudinal cohort study to evaluate novel diagnostic tools and to assess long-term impact on lung health in South African children with presumptive pulmonary TB—a study protocol. BMC Pulmonary Medicine 2023;23(1):97. doi: 10.1186/s12890-023-02329-3

