## Supplementary Appendix for "Pulmonary Function Outcomes after Tuberculosis treatment in Children: A Systematic Review and Meta-analysis"

*Online supplementary appendices*

**Appendix I**

**Search strategy**

The following PubMed search string was adapted and used for all other database searches:

| Category | Criteria | Search terms |
| --- | --- | --- |
| Population | Children under 18 years old | “child”  OR  “adolescents”  OR  “paediatric”  OR  “infant” |
| Exposure / condition | Pulmonary tuberculosis | “pulmonary tuberculosis” |
| Outcome | Lung / pulmonary involvement | “lung”  OR  “respiratory”  OR  “chest”  OR  “pulmonary” |
| Outcome | Functional / spirometry changes | “outcome”  OR  “function”  OR  “spirometry” |
| Language | English-only articles | English [Language] |

**Appendix II**

**Full details of data extraction of eligible studies**

| Included study | **Sovershaeva E, et al.^1^** | **Lee E, et al.^2^** | **Githinji LN, et al.^3^** | **Nkereuwem E, et al.^4^** | **Martinez L, et al.^5^** |
| --- | --- | --- | --- | --- | --- |
| Year Published | 2019 | 2019 | 2020 | 2022 | 2023 |
| Study design | Cross-sectional study | Cross-sectional study (retrospective) | Cohort study | Cross-sectional study | Prospective cohort (birth) |
| Date of study | 2017 Apr - 2018 Aug | 2000 Jan - 2018 Dec | 2013 Oct - 2015 Mar (enrolments) followed up at 24-months | 2020 Jan - 2021 Mar | 2012 Mar - 2015 Mar; followed up to first five years of child's life |
| Duration of study | 17 months | 18 years’ of data | 42 months | 14 months | 9 years |
| Study sites (country) | Zimbabwe | South Korea | South Africa | Gambia | South Africa |
| Study sites (details) | Harare Central Hospital paediatric HIV clinic | 28 secondary hospitals and tertiary centres across South Korea | Primary care and hospital-based ART clinics in Cape Town | Childhood TB research clinic (Medical Research Council Unit The Gambia; MRCG at LSHTM) | Births at Paarl regional hospital, TB diagnosed at TB community clinics |
| Number of children with PTB included/ total number of children in study | 57/319 | 42/341 | 305/609 | 68/159 | 95/1068 |
| Number of participants excluded due to issues with spirometry | 9 initially excluded (not part of N=319; PTB status unknown) | none, as study is retrospective | 80 with unsuccessful spirometry at 24 months (exact PTB numbers unknown) | 21 with unsuccessful spirometry at (16 post-TB and 5 controls) | 75 invalid measurements at 5 years time point (exact PTB numbers unknown) |
| Age of children in years (PTB subgroup) | Median [IQR]: 15 [12-18] | Mean (SD): 11.9 (5.6) | Mean (SD): 12 (1.6) | Median [IQR]: 8.9 [7.2-11.2] | 5 |
| Sex ratio (% males) (all participants) | 157:162 (49.2% males) | 207:180 (53.5% males) | 309:300 (50.7% males) | 93:66 (58.5% males) | 549:519 (51.5% males) |
| Primary diagnostic method (number of bacteriologically confirmed cases/all TB cases in study) | Unspecified | Extracted from hospital records. 59.5% with Mtb confirmed (25/42) | Extracted from hospital records and validated study questionnaires | Positive culture, Xpert MTB/RIF, or both from at least one respiratory specimen. 35.3% confirmed TB (24/68) | Positive culture, Xpert MTB/RIF, or AFB smear test. 13.7% microbiological confirmation from all PTB cases (13/95) |
| Other diagnostic methods  (if specified) | GeneXpert Ultra MTB/RIF used to rule out active TB | Mantoux skin testing (14 positives from 80 tests done) | Unspecified | At least two of: a) symptoms and signs suggestive of TB, b) CXR consistent with TB, c) close TB exposure, and d) positive response to TB treatment 64.7% (44/68) of cases with unconfirmed TB | At least two of: a) CXR suggestive with TB, b) close TB exposure, or c) positive tuberculin skin test, signs and symptoms of TB. 37.5% had suggestive CXR (36/96); 5% (5/96) with 2 episodes of TB |
| Baseline lung function (all participants) | Unspecified | Unspecified | FEV1 z-score of -1.0 (SD=1.3) for HIV-positive sub-group, 60% had past history of TB | Unspecified | Performed at 6 weeks after birth, at 1 year of age, then annually until 5 years old |
| Details of other baseline pulmonary investigations  (all participants) | atopy (asthma, eczema, etc) present in 21% (12/57) | Chronic suppurative cough, pneumonia, wheezing, haemoptysis, etc., present in 68% (244/359) | Data collected for asthma history and smoke exposure (passive and active) | Previous asthma (n=3); non-TB LRTI more common in post-TB cases (6/68, 8.8%) than non-TB group (1/91, 1.1%) in preceding year | Baseline wheezing assessment carried out at 6 weeks |
| Other pulmonary investigations outside baseline  (all participants) | eNO testing was done before PFT | Unspecified | Data collected for nontuberculous LRTI, asthma, cough, wheeze, and shortness of breath | unspecified | Routine investigations done during each year of life |
| Risk factors: Primary immunodeficiency | Unspecified | Unspecified | Unspecified | Unspecified | Unspecified |
| Risk factors: Pre-term birth (all participants) | Unspecified | 21/387 born at less than 37 weeks gestation | Unspecified | Unspecified | 21.9% (21/96) among TB cases compared to 15.2% (148/972) |
| Risk factors: Others | Unspecified | Unspecified | Unspecified | Unspecified | Low birth weight (<2.5kg) found in 18.8% of TB cases (18/96) compared to 13.0% (139/972) for non-TB |
| Inclusion criteria | 1) 6-19 years old, 2) perinatally acquired HIV, 3) Been on ART for at least 6 months | 1) Bronchiectasis diagnosed according to ICD-10 code, 2) CT-confirmed bronchiectasis by two independent paediatric respiratory specialists based on broncho-arterial ratio >1:1 | 1) 9-14 years old, 2) vertically transmitted HIV, 3) ART for at least 6 months, 4) awareness of HIV status, and 5) consent and assent obtained | 1) 15 years old or younger at diagnosis, 2) successfully completed anti-TB treatment with documented outcome of "cured" or "completed" at least 6 months prior to enrolment | 1) Pregnant mothers within catchment area of main study, 2) children with a) ≥1 valid lung function or anthropometric measurement, or b) evaluated for wheezing through 5 years of age. |
| Exclusion criteria | Active TB or acute respiratory tract infection at enrolment | NA (did not fulfil inclusion criteria) | NA (did not fulfil inclusion criteria) | 1) Younger than 5 years old, 2) unwilling to participate, 3) relocated, or 4) recurrent PTB | 1) Expectant mothers: a) under 18 years old, b) plan to move away within 1 yr, c) lost to antenatal follow-up, or d) pregnancy loss; 2) Children who lacked relevant measures |
| Comorbidity: HIV  (all participants) | TB sub-group were all HIV-positive (100%) | Unspecified | 303 HIV-positives /305 total number with TB | 13.2% (9/68) in post-TB; none (0/91) in controls | Overall study cohort had 22.1% (236/1068 cases) exposed to maternal HIV but remained uninfected, only 2 infants (0.2%) living with HIV |
| Comorbidity: Malnutrition | TB-subgroup showed evidence of wasting (52.6%) and stunting (45.6%) | Overall study cohort had 18/341 cases of growth retardation.  (exact figure for PTB subgroup unknown) | Overall study cohort had mean BMI of 17.7 (SD: 3.2) at baseline.  (exact figure for PTB subgroup unknown) | In post-TB cases, 25% (17/68) underweight; 19.1% (13/68) stunted | significant difference in Length-for-age, Weight-for-age z-scores (p<0.05), lower in TB, compared to no TB between 1 to 4 years of age |
| Comorbidity: Others | Unspecified | Abnormal heart function, secondary pulmonary hypertension, decreased renal function and abnormal liver function was assessed for overall study cohort | Unspecified | Unspecified | Maternal antenatal smoking in23.3% of total cases (247/1068) |
| Setting: LMIC | Yes. | No. | No. | Yes | No |
| Setting: National HIV-prevalence (children under 15 years old) | ZW: 0.015% | Unavailable | ZA: 0.021% | GM: 0.0014% | ZA: 0.021% |
| Set: Rural / urban / other (specify) | Urban | Urban | Urban | Urban | Mixed |
| Concurrent medication | All HIV-positive participants were on ART; up to 43.9% on protease inhibitors | Unspecified | All HIV-positive participants were on ART; some were on corticosteroids, salbutamol, steroids, as well as cotrimoxazole and isoniazid prophylaxis | Unspecified | Unspecified |
| % EPTB involvement | Unspecified.  Recall bias likely. | Unspecified | Unspecified | Unspecified | 1 case of tuberculous meningitis (not PTB) |
| % drug-resistant TB | Unspecified | Unspecified | Unspecified | Unspecified | 1 case of MDR-TB |
| % completed treatment | Unspecified; presumably 100% as all were negative on Xpert MTB/RIF | Unspecified; presumably 100% completed treatment due to strict national guidelines | Unspecified; Presumably all. | 100% of included | 100% of included |
| Time elapsed since completing anti-TB treatment (months) | Unspecified | Unspecified | ≥24 months | 19.2 (IQR: 10.2-44.4) | At least one year after TB diagnosis, or 6 months from end of treatment. |
| Treatment regimen | Unspecified | Unspecified | Unspecified | Unspecified | 2HRZ(E)/4HR according to national guidelines, with modifications made if drug-resistant |
| Spirometry results: FEV_1_ | z-score, Median [IQR]:  Cases: -1.8 [-2.3 to -1.3]  Controls: -1.1 [-1.3 to 0.4] | Mean % predicted:  69.9% (SD not numerically reported) | z-score, coeff. (95% CI):  Cases: -0.27 (-0.50 to -0.03)  Control: -0.12 (-0.46 to 0.23) | z-score, Mean (SD): Cases: -1.52 (0.99)  Controls: -0.83 (0.84) | Not done |
| Spirometry results: FVC | Not done | Mean % predicted:  74.0% (SD not numerically reported) | z-score, coeff. (95% CI):  Cases: -0.27 (-0.49 to -0.05)  Controls: 0.01 (-0.31-0.33) | z-score, Mean (SD): Cases: -1.32 (1.02) Controls: -0.87 (0.89) | Not done |
| Spirometry results: FEV_1_/FVC | Not done | Not done | z-score, coeff. (95% CI):  -0.01 (CI not reported) | z-score, Mean (SD):  Cases: -0.54 (0.91) Controls: -0.03 (0.81) | Not done |
| Other lung function tests | Not done | Not done | Not done | Not done | Documented in Appendix VI |
| Calculated/derived measures | Assuming normality, FEV1 z-score **mean** (SD) = -1.8 (0.74) | rspiro-derived (GLI-2012) FEV1 z-score = -3.26 (SD 0.81); FVC z-score = -3.18 (SD 0.72) | Not applicable | Not applicable | Not applicable |
| SO: Other lung function indices | Not applicable | Not applicable | Not applicable | Spirometry pattern: 36.4% (19/52) restrictive, 1.9% (1/52) obstructive | Not applicable |
| Funding sources | This study was supported by the Global Health and Vaccination Research (GLOBVAC) Programme of the Medical Research Council of Norway and by HelseNord (HNF 1387-17), and the Wellcome Trust (206316/Z/17/Z). | This research did not receive any specific grant from funding agencies in the public, commercial, or not-for-profit sectors. | Funded by National Institute of Child Health and Human Development (grant number R01HD074051), the South Africa Medical Research Council, and the African Partnership for Chronic Diseases Research. | Funded by an Imperial College London Wellcome Trust Institutional Strategic Support Fund (grant no PS3456_WMNP), the UKRI-GCRF, and MRC (programme grants MR/P024270/1 and MR/K011944/1). The funder had no role in study design, data collection, data analysis, data interpretation or writing. | Funded by the Bill & Melinda Gates Foundation (grant number OPP 1017641), Medical Research Council South Africa, National Research Foundation South Africa, the National Institutes of Health H3 Africa (grant numbers U54HG009824, U01AI110466), and the Wellcome Trust (grant numbers, 098479/Z/12/Z and 204755/Z/16/Z) |

**Appendix III**

**Characteristic of excluded studies**

During initial title and abstract screening, about half the records were excluded due to wrong study design, i.e., animal models, diagnostic studies, vaccine trials, etc. Studies were also rejected due to non-children populations, TB without pulmonary involvement, foreign language studies with English abstracts, non-PFT treatment outcomes.

Several studies appeared to have met inclusion criteria were excluded after full text review revealed adult-only populations not apparent initially,^6-14^ mixed adolescent-and-adult populations with insufficient data for the under-18 years old group^15-18^, and studies which did not perform PFTs as a reported outcome^19 20^. One excluded study^21^ described the baseline for a longitudinal cohort, which overlapped with an included study which reported lung function decline associated with PTB at the 24-month endpoint.^3^

**Appendix IV**

**Newcastle Ottawa Scale for Study Quality and Risk of Bias assessment**

| **Included studies** | Sovershaeva *et al*^1^ | Lee *et al*^2^ | Githinji *et al*^3^ | Nkereuwem *et al*^4^ | Martinez *et al*^5^ |
| --- | --- | --- | --- | --- | --- |
| **Selection** |  |  |  |  |  |
| ***1) Representativeness of the exposed cohort***   a) truly representative of average child with PTB within community * b) somewhat representative of average child with PTB * c) indications of non-random sampling, e.g. only hospitalised cases, or common history of other non-PTB disease d) no description of the derivation of the cohort | **0** | **0** | **0** | **1** | **1** |
| ***2) Selection of the non-exposed cohort***   a) drawn from the same community as the exposed cohort * b) drawn from a different community c) no description of the derivation of the non-exposed cohort | **1** | **0** | **1** | **1** | **1** |
| ***3) Ascertainment of exposure***   a) bacteriological confirmation of TB * b) secure record (e.g. medical/lab records) * c) written self-report d) no description | **0** | **1** | **1** | **1** | **1** |
| ***4) Demonstration that lung function changes due to PTB was not present at start of study***   a) yes * b) no | **0** | **0** | **0** | **0** | **1** |
| **Comparability (up to 2 *)** |  |  |  |  |  |
| ***1) Comparability of cohorts on the basis of the design or analysis***   a) study controls for height and age (or height-to-age) * b) study controls for sex, and (past LRTI OR other respiratory disorders OR immune-deficiencies for non-HIV cohorts) * | **2** | **1** | **2** | **2** | **2** |
| **Outcome** |  |  |  |  |  |
| ***1) Assessment of outcome***   a) valid spirometry * b) valid non-spirometry pulmonary function tests * c) self-reported d) no description | **1** | **1** | **1** | **1** | **1** |
| ***2) Same method of ascertainment for cases and controls***   a) yes * b) no | **1** | **1** | **1** | **1** | **1** |
| ***3) Adequacy of follow up of cohorts***  ***NOT APPLICABLE FOR CROSS-SECTIONAL STUDIES***   a) complete follow up - all subjects accounted for * b) subjects lost to follow up no more than 10% of surviving * c) follow up rate <90% of surviving, insufficient description of loss d) no statement | NA | NA | **1** | NA | **1** |
| **Quality Score** | **5/8** | **4/8** | **7/9** | **7/8** | **9/9** |

**Appendix V**

**Additional details of included studies**

Sovershaeva *et al*^1^ reported median FEV_1_ z-score as part of the BREATHE trial,^22^ exhaled nitric oxide was measured as primary study endpoint, z-score for FEV_1_ was reported but not FVC. History of TB was determined via self-reporting, thus limited by recollection bias.

Lee *et al*^2^ reviewed hospital records for PTB diagnosis and spirometry results, PFT findings may not be solely attributed to PTB as within-study subgroups were classified by primary aetiologies and did not preclude co-contributors. Mean percentage of predicted FEV_1_ and FVC were reported without numerical values for summary statistics, but an overall negative effect of TB and other aetiologies of bronchiectasis on lung function of South Korea children was clearly observed. While percentage of predicted FEV_1_/FVC ratio was plotted, the numerical value of the mean was incorrect, attempts to contact study authors for correct data were futile.

Githinji *et al*^3^ reported association coefficients for the effect of PTB on FEV_1_, FVC, FEV_1_/FVC z-scores carried out as part of a larger Cape Town Adolescent Antiretroviral Cohort (CTAAC) study^21^ with nearly half the cohort with perinatally-infected HIV having history of TB.

Nkereuwem *et al*^4^ performed spirometry on the youngest cohort among studies included in meta-analyses, and reported FEV_1_, FVC, FEV_1_/FVC z-scores less likely to be confounded by other diseases, with low rate of HIV-coinfections (13.2%) indicative that changes in lung function measures were less likely to be biased by other non-PTB factors.

Martinez *et al*^5^ reported longitudinal pulmonary changes due to PTB using non-spirometry PFTs (p<0.05).

**Appendix VI**

**Association coefficients from included study which did not perform spirometry as PFT of choice (Tuberculosis versus no Tuberculosis between 1-4 years of age).^5^**

| Lung function measurements | N *^a^* | Coefficient | (95% CI), | P-value |
| --- | --- | --- | --- | --- |
| Functional residual capacity (L) | 993 | -0.01 | (-0.02, 0.01) | 0.723 |
| Lung clearance index (n turnovers) | 993 | -0.06 | (-0.21, 0.09) | 0.281 |
| Tidal volume (mL) | 997 | -9.32 | (-14.89, -3.75) | 0.004 ** |
| Respiratory rate (breaths min^-1^) | 997 | 1.52 | (-0.35, 3.40) | 0.109 |
| Minute ventilation | 997 | -67.86 | (-237.44, 101.72) | 0.705 |
| t_PTEF_/t_E_ (%) | 997 | -2.73 | (-5.45, -0.01) | 0.049 * |
| t_I_/t_TOT_ (%) | 997 | 0.10 | (-0.79, 0.99) | 0.981 |
| Compliance (mL hPa^–1^) | 962 | -0.001 | (-0.002, 0.000) | 0.208 |
| Resistance (hPa sL^–1^) | 962 | 0.58 | (1.82, 3.00) | 0.637 |

*^a^* N = number of participants with valid lung function values at 5 years or older, from which up to 9.9% (*n=95*) suffered from childhood PTB.

**References**

1. Sovershaeva E, Kranzer K, McHugh G, et al. History of tuberculosis is associated with lower exhaled nitric oxide levels in HIV-infected children. *Aids* 2019;33(11):1711-18. doi: 10.1097/qad.0000000000002265 [published Online First: 2019/05/21]

2. Lee E, Shim JY, Kim HY, et al. Clinical characteristics and etiologies of bronchiectasis in Korean children: A multicenter retrospective study. *Respir Med* 2019;150:8-14. doi: 10.1016/j.rmed.2019.01.018 [published Online First: 2019/04/10]

3. Githinji LN GD, Hlengwa S, Machemedze T, Zar HJ. Longitudinal Changes in Spirometry in South African Adolescents Perinatally Infected With Human Immunodeficiency Virus Who Are Receiving Antiretroviral Therapy. *Clinical infectious diseases : an official publication of the Infectious Diseases Society of America* 2020;70(3):483-90. doi: doi:

4. Nkereuwem E, Agbla S, Sallahdeen A, et al. Reduced lung function and health-related quality of life after treatment for pulmonary tuberculosis in Gambian children: a cross-sectional comparative study. *Thorax* 2023;78(3):281-87. doi: 10.1136/thorax-2022-219085

5. Martinez L, Gray DM, Botha M, et al. The Long-Term Impact of Early-Life Tuberculosis Disease on Child Health: A Prospective Birth Cohort Study. *Am J Respir Crit Care Med* 2023;0(ja):null. doi: 10.1164/rccm.202208-1543OC [published Online First: 2023/02/07]

6. Lee SW, Kim YS, Kim DS, et al. The risk of obstructive lung disease by previous pulmonary tuberculosis in a country with intermediate burden of tuberculosis. *J Korean Med Sci* 2011;26(2):268-73. doi: 10.3346/jkms.2011.26.2.268 [published Online First: 2011/02/03]

7. Nihues SdSE, Mancuzo EV, Sulmonetti N, et al. Chronic symptoms and pulmonary dysfunction in post-tuberculosis Brazilian patients. *The Brazilian Journal of Infectious Diseases* 2015;19(5):492-97. doi: <https://doi.org/10.1016/j.bjid.2015.06.005>

8. Cole G, Miller D, Ebrahim T, et al. Pulmonary impairment after tuberculosis in a South African population. *S Afr J Physiother* 2016;72(1):307. doi: 10.4102/sajp.v72i1.307 [published Online First: 2016/06/30]

9. Manji M, Shayo G, Mamuya S, et al. Lung functions among patients with pulmonary tuberculosis in Dar es Salaam - a cross-sectional study. *BMC Pulm Med* 2016;16(1):58. doi: 10.1186/s12890-016-0213-5 [published Online First: 2016/04/25]

10. Fiogbe AA, Agodokpessi G, Tessier JF, et al. Prevalence of lung function impairment in cured pulmonary tuberculosis patients in Cotonou, Benin. *Int J Tuberc Lung Dis* 2019;23(2):195-202. doi: 10.5588/ijtld.18.0234 [published Online First: 2019/02/28]

11. Hanekom S, Pharaoh H, Irusen E, et al. Post-tuberculosis health-related quality of life, lung function and exercise capacity in a cured pulmonary tuberculosis population in the Breede Valley District, South Africa. *South African Journal of Physiotherapy* 2019;75(1):1-8.

12. Osman M, Welte A, Dunbar R, et al. Morbidity and mortality up to 5 years post tuberculosis treatment in South Africa: A pilot study. *Int J Infect Dis* 2019;85:57-63. doi: 10.1016/j.ijid.2019.05.024 [published Online First: 2019/05/28]

13. Ojuawo OB, Fawibe AE, Desalu OO, et al. Spirometric abnormalities following treatment for pulmonary tuberculosis in Ilorin, Nigeria. *Niger Postgrad Med J* 2020;27(3):163-70. doi: 10.4103/npmj.npmj_18_20 [published Online First: 2020/07/21]

14. Tadolini M, Codecasa LR, García-García JM, et al. Active tuberculosis, sequelae and COVID-19 co-infection: first cohort of 49 cases. *Eur Respir J* 2020;56(1) doi: 10.1183/13993003.01398-2020 [published Online First: 2020/05/28]

15. Singla N, Singla R, Fernandes S, et al. Post treatment sequelae of multi-drug resistant tuberculosis patients. *Indian J Tuberc* 2009;56(4):206-12. [published Online First: 2010/05/18]

16. Amorim E, Saad R, Jr., Stirbulov R. Spirometry evaluation in patient with tuberculosis sequelae treated by lobectomy. *Rev Col Bras Cir* 2013;40(2):117-20. doi: 10.1590/s0100-69912013000200006 [published Online First: 2013/06/12]

17. Ko JM, Park HJ, Cho DG, et al. CT differentiation of tuberculous and non-tuberculous pleural infection, with emphasis on pulmonary changes. *Int J Tuberc Lung Dis* 2015;19(11):1361-8. doi: 10.5588/ijtld.15.0092 [published Online First: 2015/10/16]

18. Gandhi K, Gupta S, Singla R. Risk factors associated with development of pulmonary impairment after tuberculosis. *Indian J Tuberc* 2016;63(1):34-8. doi: 10.1016/j.ijtb.2016.01.006 [published Online First: 2016/05/29]

19. Apis V, Landi M, Graham SM, et al. Outcomes in children treated for tuberculosis with the new dispersible fixed-dose combinations in Port Moresby. *Public Health Action* 2019;9(Suppl 1):S32-s37. doi: 10.5588/pha.18.0062 [published Online First: 2019/10/04]

20. Gafar F, Van't Boveneind-Vrubleuskaya N, Akkerman OW, et al. Nationwide analysis of treatment outcomes in children and adolescents routinely treated for tuberculosis in the Netherlands. *Eur Respir J* 2019;54(6) doi: 10.1183/13993003.01402-2019 [published Online First: 2019/09/14]

21. Githinji LN GD, Hlengwa S, Myer L, Zar HJ. Lung Function in South African Adolescents Infected Perinatally with HIV and Treated Long-Term with Antiretroviral Therapy. *Annals of the American Thoracic Society* 2017;14(5):722-29. doi: doi:

22. Gonzalez-Martinez C, Kranzer K, McHugh G, et al. Azithromycin versus placebo for the treatment of HIV-associated chronic lung disease in children and adolescents (BREATHE trial): study protocol for a randomised controlled trial. *Trials* 2017;18(1):622. doi: 10.1186/s13063-017-2344-2
